# Differential WMH progression trajectories in progressive and stable mild cognitive impairment

**DOI:** 10.1101/2022.09.21.22280209

**Authors:** Farooq Kamal, Cassandra Morrison, Josefina Maranzano, Yashar Zeighami, Mahsa Dadar, Alzheimer’s Disease Neuroimaging Initiative

## Abstract

**Background:** Pathological brain changes such as white matter hyperintensities (WMHs) occur with increased age and contribute to cognitive decline. Current research is still unclear regarding the association of amyloid positivity with WMH burden and progression to dementia in people with mild cognitive impairment (MCI).

**Methods:** This study examined whether WMH burden increases differently in both amyloid-negative (Aβ-) and amyloid-positive (Aβ+) people with MCI who either remain stable or progress to dementia. We also examined regional WMHs differences in all groups: amyloid positive (Aβ+) progressor, amyloid negative (Aβ–) progressor, amyloid positive (Aβ+) stable, and amyloid negative (Aβ–) stable. MCI participants from the Alzheimer’s Disease Neuroimaging Initiative were included if they had APOE ɛ4 status and if they had amyloid measures to determine amyloid status (i.e., positive, or negative). A total of 820 MCI participants that had APOE ɛ4 status and amyloid measures were included in the study with 5054 follow-up time points over a maximum period of 13 years with an average of 5.7 follow-up timepoints per participant. Linear mixed-effects models were used to examine group differences in global and regional WMHs.

**Results:** People who were Aß– stable had lower baseline WMHs compared to both Aß+ progressors and Aß+ stable across all regions. When examining change over time, compared to Aß– stable, all groups had steeper change in WMH burden with Aß+ progressors having the largest change (largest increase in WMH burden over time).

**Conclusion:** These findings suggest that WMH progression is a contributing factor to conversion to dementia both in amyloid-positive and negative people with MCI.

## 1 Introduction

Mild cognitive impairment (MCI) is characterized by declines in cognitive functioning that go beyond what is observed in normal aging, but do not interfere with activities of daily living (Petersen et al., 2014). People with MCI experience both an increased rate of cognitive decline and progress to dementia with a higher annual conversion rate compared to healthy older adults (Petersen, 2016; Petersen et al., 2014). While not everyone with MCI progresses to dementia, this stage of cognitive decline is often identified as a transitional stage between healthy aging and dementia. Much research has thus focused on studying people with MCI to identify who will eventually convert to dementia. More specifically, researchers endeavor to find early biomarkers in people with MCI who convert to dementia (hereafter referred to as progressive MCI, pMCI) that distinguish them from people with MCI who do not convert to dementia (hereafter referred to as stable MCI, sMCI). Identifying who will progress from MCI to dementia has been difficult in clinical practice because of the heterogeneous nature of MCI. However, one approach is to examine people with MCI who have biomarkers that increase their risk of developing dementia. For example, people with MCI who are also positive for beta-amyloid (Aβ) or pathologic tau, key early events in the pathophysiological process of Alzheimer’s disease (AD), are more likely to progress to AD and may be in the earliest symptomatic stages of AD (see Sperling et al., 2014 for a review).

Other pathological brain changes, such as white matter hyperintensities (WMHs), have been shown to contribute to healthy older adults’ risk for cognitive decline (Morrison et al., 2022) and progression to MCI or dementia (Bangen et al., 2018; Kim et al., 2015; Yoshita et al., 2006). WMHs are observed as increased signal in T2-weighted or fluid-attenuated inversion recovery (FLAIR) magnetic resonance images (MRIs). WMHs are used as a proxy for cerebrovascular disease, a known contributor for cognitive decline and dementia (Abraham et al., 2016; Tamura & Araki, 2015; Van Der Flier et al., 2018). High WMH burden is associated with increased cognitive decline in MCI (Hirao et al., 2021; Kamal et al., 2022; Kim et al., 2015; Li et al., 2016) and increased rate of progression from MCI to dementia (Dadar et al., 2019). When observing pMCI in Parkinson’s disease (PD) dementia, people with PD-MCI who progressed to Parkinson’s disease dementia, at least 24 months later, had higher WMH volumes compared to PD-sMCI (Sunwoo et al., 2014). One study found that over an 18-month period, pMCI was associated with increased periventricular and deep WMHs compared to sMCI (Prasad et al., 2011). However, the sample size in this study was quite limited. In a much larger sample of 591 people with MCI, Dadar et al., (2019) observed that those with pMCI had increases in total WMH volume compared to sMCI (with mean follow-up times of 2-2.5 years). While these studies offer insight into the association between WMHs and MCI, the follow-up periods are quite short given that the annual conversion rate from MCI to dementia in community and clinic samples is only 3% and 13%, respectively (Farias et al., 2009).

Several other limitations exist in the current research examining the relationship between WMHs and progressive vs. stable MCI. These studies used a total WMH approach which captures overall lesion volume affecting the whole brain. Examining regional WMHs is important when examining conversion from MCI to dementia because different patterns of WMH accumulation are associated with different types of dementia. For example, a more widespread WMH accumulation in people with MCI is associated with progression to vascular dementia (Bombois et al., 2008) whereas posterior WMHs are associated with AD (Brickman, 2013; Habes et al., 2018). These findings suggest that the evaluation of regional WMH differences may provide insight into the type of dementia someone with MCI will develop. Furthermore, previous studies have not examined WMH differences between pMCI and sMCI who are Aβ+ and Aβ–. Those with MCI who are Aβ+ are on the AD trajectory, whereas Aβ– are more likely to develop other types of dementia. This difference in diagnostic outcomes may also influence WMH burden. Previous research examining amyloid positivity in mild amnestic type dementia found that those who are Aβ+ have increased WMHs compared to those who are Aβ–. Ultimately, while previous studies have observed that people with pMCI have increased WMH burden compared to sMCI, they have limited sample size, short follow-up times, have not examined WMH burden in a regional approach, and have not compared Aβ+ and Aβ– participants. More research is therefore needed to understand the association of amyloid positivity with WMH burden and progression to dementia in people with MCI.

Using data from the Alzheimer’s Disease Neuroimaging Initiative (ADNI), the present study was designed to expand on the current research by examining differences in WMH burden between Aβ+ and Aβ– progressive and stable MCI participants over a follow-up period of 13 years (820 participants, with an average of 5.7 follow-up timepoints per participant). The goal of this study was to examine whether WMHs influence conversion to dementia in both amyloid negative and amyloid positive people with MCI. Additionally, we sought to examine how much of WMH progression is associated with amyloid positivity vs. vascular risk factors by including vascular risk factors into our models. This distinction between amyloid vs. vascular causes of WMHs is essential since many vascular risk factors associated with WMHs might be potentially treatable or preventable (Scott et al., 2015; Snyder et al., 2015; Yamada & Naiki, 2012). Therefore, if we observe that WMHs in these groups are associated with the vascular risk factors, then progression to dementia may be mitigated or prevented by treating these underlying risk factors.

## 2 Methods

### 2.1 Alzheimer’s Disease Neuroimaging Initiative

The data used in the preparation of this article were obtained from the ADNI database (adni.loni.usc.edu). The ADNI was launched in 2003 as a public-private partnership, led by Principal Investigator Michael W. Weiner, MD. The primary goal of ADNI has been to test whether serial magnetic resonance imaging (MRI), positron emission tomography (PET), other biological markers, and clinical and neuropsychological assessment can be combined to measure the progression of mild cognitive impairment (MCI) and early Alzheimer’s disease (AD). The study received ethical approval from the review boards of all participating institutions. Written informed consent was obtained from participants or their study partner. Participants were included from all ADNI cohorts (ADNI-1, ADNI-2, ADNI-GO, and ADNI-3).

### 2.2 Participants

Participant inclusion and exclusion criteria are available at www.adni-info.org. All participants were between 55 and 90 years of age at the time of recruitment and exhibited no evidence of depression. MCI participants obtained scores between 24 and 30 on the Mini-Mental Status Examination (MMSE), 0.5 on the Clinical Dementia Rating (CDR), and exhibited abnormal scores on the Wechsler Memory Scale. Participants were only included if they had a reported Apolipoprotein E (APOE) ɛ4 status and if they had amyloid measures to determine amyloid status (i.e., positive, or negative). A total of 820 MCI participants had both APOE ɛ4 status and amyloid measures and were thus included in the study with 5054 follow-up time points over a maximum period of 13 years with an average of 5.7 follow-up timepoints per participant.

MCI participants were divided into four groups: 1) amyloid positive (Aβ+) progressor, 2) amyloid negative (Aβ–) progressor, 3) amyloid positive (Aβ+) stable, 4) amyloid negative (Aβ–) stable. Both PET and CSF values were used to determine amyloid positivity in people with MCI because some participants had only one measurement available. Amyloid positivity was identified based on participants meeting one of the following criteria: 1) a standardized uptake value ratio (SUVR) of > 1.11 on AV45 PET (Landau et al., 2013), or 2) an SUVR of >1.2 using Pittsburgh compound-B PET (Villeneuve et al., 2015), or 3) an SUVR of ≥1.08 for Florbetaben (FBB) PET (S. Landau et al., 2011), or 4) a cerebrospinal fluid Aß1-42 ≤ 980 pg/ml as per ADNI recommendations. Participants were considered as a progressor if they had a diagnosis of dementia on their last follow-up visit and considered as stable if they still had a diagnosis of MCI on their last follow-up visit. The groups had the following number of participants: 1) Aß+ Progressor: 239 with 1685 follow-ups (average follow-up times of 7.05 years), 2) Aß– Progressor: 22 with 140 follow-ups (average follow-up times of 6.36 years), 3) Aß+ stable: 343 with 1984 follow-ups (average follow-up times of 5.65 years), and 4) Aß– Stable: 216 with 1291 follow-ups (average follow-up times of 5.98 years). Only participants that had multiple follow-up visits were included in the study. To ensure that the minimum duration of follow-up time does not impact our study findings, the models were repeated including participants with a minimum of one, two, and three years of follow-up.

### 2.3 Structural MRI acquisition and processing

Standardized acquisition protocols designed and implemented by ADNI were used for scanning of all participants. Detailed MRI protocols and imaging parameters can be found at: http://adni.loni.usc.edu/methods/mri-tool/mri-analysis/. All participant MRI data (baseline and longitudinal) were downloaded from the ADNI public website.

All T1w scans were pre-processed using our standard pipeline which includes: noise reduction (Coupe et al., 2008), intensity inhomogeneity correction (Sled et al., 1998), and intensity normalization into range [0-100]. These pre-processed images were then linearly (9 parameters: 3 translation, 3 rotation, and 3 scaling) (Dadar, Fonov, et al., 2018) registered to the MNI-ICBM152-2009c average template (Fonov et al., 2011).

### 2.4 WMH measurements

A previously validated WMH segmentation technique that has been extensively tested for assessment of WMHs in aging and neurodegenerative disorders was used to obtain WMH measurements (Dadar et al., 2017). This technique has been employed in other multi-center studies (Anor et al., 2021; Dadar et al., 2020) and has also been validated in the ADNI cohort (Dadar et al., 2019). Automatic segmentation of the WMHs was completed using the T1w contrasts, along with location and intensity features from a library of manually segmented scans (50 ADNI participants independent of the ones studied here) in combination with a random forest classifier to detect the WMHs in new images (Dadar et al., 2017). The WMHs were segmented using T1w images (instead of FLAIR and T2w/PD scans) since ADNI1 only acquired T2w/PD images with resolutions of 1×1×3 mm^3^, whereas ADNI2/GO acquired only axial FLAIR images with resolutions of 1×1×5 mm^3^, and ADNI3 acquired sagittal FLAIR images with resolutions of 1×1×1.2 mm^3^. Hence, since these inconsistencies might have made direct comparisons of WMH measurements across ADNI1 and ADNI2/3/GO studies unreliable, we opted to use T1w images that were consistently acquired to estimate WMH burden. Further, we have previously demonstrated that these T1w-based WMH volumes are very highly correlated with FLAIR and T2w based WMH loads in the ADNI dataset (Dadar, Maranzano, et al., 2018). The quality of the registrations and WMH segmentations was visually assessed (by M.D.) and the cases that did not pass this quality control step were excluded from the analyses (N = 59). WMH load was defined as the volume of all voxels identified as WMH in the standard space (in mm^3^) and were thus normalized for head size. Regional and total WMH volumes were calculated based on Hammers Atlas (Dadar et al., 2017; Dadar, Maranzano, et al., 2018). All WMH volumes were also log-transformed to achieve normal distribution.

### 2.6 Statistical Analysis

#### 2.6.1 Baseline Assessments

Participant demographic information is presented in Table 1. T-tests and chi-square analyses were performed on the demographic information and corrected for multiple comparisons using Bonferroni correction. The following linear regression models were used to investigate group differences in total and regional WMH loads, including age, sex, years of education, and APOE4 status as covariates. The categorical variable of interest was group status (Aß+ Progressor, Aß– Progressor, Aß+ Stable, and Aß– Stable) to examine whether group status influenced baseline WMH.

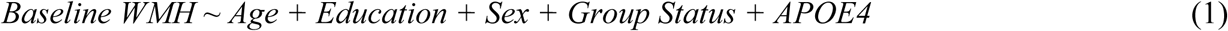

**Table 1:** Demographic and clinical characteristics for Aß+ Progressor, Aß– Progressor, Aß+ Stable, and Aß– Stable

| Demographic Information | Aβ+ Progressor<br>n = 239 | Aβ- Progressor<br>n = 22 | Aβ+ Stable<br>n = 343 | Aβ- Stable<br>n = 216 |
| --- | --- | --- | --- | --- |
| Baseline Age | 73.4 ± 6.6 | 74.9 ± 7.0 | 73.7 ± 7.3 | 70.9 ± 8.2<sup>1</sup> |
| Education | 15.8 ± 2.8 | 16.5 ± 2.2 | 16.1 ± 2.8 | 16.0 ± 2.7 |
| APOE ε4+ | 165 (69%)<sup>2</sup> | 2 (9%)<sup>2</sup> | 186 (54%)<sup>2</sup> | 38(18%)<sup>2</sup> |
| Male sex | 136 (57%) | 16 (73%) | 208 (60%) | 125 (58%) |
| ADAS-13 | 19.8 ± 6.8<sup>3</sup> | 16.8 ± 5.57<sup>3</sup> | 14.6 ± 5.9<sup>3</sup> | 11.9 ± 5.2<sup>3</sup> |
| CDR-SB | 1.76 ± 1.03<sup>4</sup> | 1.52 ± 1.06<sup>4</sup> | 1.14 ± 0.83<sup>4</sup> | 1.10 ± 0.74<sup>4</sup> |
| BMI | 26.3 ± 4.30 | 28.9 ± 5.24 | 27.2 ± 4.83 | 27.8 ± 4.59<sup>5</sup> |
| Hypertension | 101 (42%) | 13 (60%) | 176 (51%) | 105 (49%) |
| Diabetes | 17 (.07%) | 3 (14%) | 30 (8%) | 19 (9%) |
| BP Systolic | 133 ± 18.5 | 135 ± 17.5 | 133 ± 19.2 | 133 ± 15.4 |
| BP Diastolic | 73.5 ± 10.7 | 73.6 ± 10.3 | 75.0 ± 9.88 | 74.3 ± 9.77 |
| Median Baseline WMH | 6011 (2897 – 50650) | 6219 (3360 – 26101) | 6202 (2146 – 50911) | 4768 (2762 – 36006) |

Notes: values are expressed as mean ± standard deviation, or number of participants (percentage %). Significance reported if result survived correction of multiple comparison. <sup>1</sup> = Aβ- Stable was significantly younger than both Aβ+ Stable and Aβ+ Progressor. <sup>2</sup> = The proportion of people with APOE ε4+ significantly differed between all groups except Aβ- Stable and Aβ- Progressor. <sup>3</sup> = All groups significantly differed except for both Aβ- and Aβ+ Progressors, and Aβ- Progressor and Aβ+ Stable. <sup>4</sup> = Aβ+ Progressor significantly differed from Aβ+ Stable and Aβ- Stable <sup>5</sup> = Aβ+ Progressor and Aβ- Stable significantly differed. APOE ε4+ = number and percentage of people with at least one ε4 allele. ADAS-13 = Alzheimer's Disease Assessment Scale -13. CDR-SB = Clinical Dementia Rating – Sum of Boxes. BMI = body mass index. BP = blood pressure. WMH = white matter hyperintensity in mm<sup>3</sup>

To investigate the impact of presence of vascular risk factors on WMH burden, the same analysis was completed a second time including hypertension, diabetes status, BMI, diastolic and systolic blood pressure as additional covariates. History of hypertension was a categorical factor whereas measures of systolic and diastolic blood pressure were continuous.

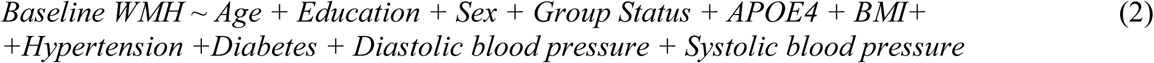

#### 2.6.3 Longitudinal Assessments

Differences in WMH progression between Aß+ pMCI, Aß– pMCI, Aß+ sMCI, and Aß– sMCI were investigated using linear mixed effects models. To do so, we examined WMH load (frontal, temporal, parietal, occipital, and total) at different visits as the dependent variable of interest and the interaction between follow-up time and group status (i.e., Aß+ Progressor, Aß– Progressor, Aß+ Stable, and Aß– Stable) as the main dependent variables. Regional WMH values (i.e., frontal, temporal, parietal, and occipital) were summed across the right and left hemispheres to obtain one measure for each region. All results were corrected for multiple comparisons using false discovery rate (FDR), p-values are reported as raw values with significance then determined by FDR correction (Benjamini et al., 1995).

The main variable of interest was the interaction term between follow-up time and group status based on group status (i.e., Aß+ or Aß-, Progressor or Stable), contrasting the slopes of changes in WMH burden for MCI: Aß+ Progressor, Aß– Progressor, and Aß+ Stable against the Aß– Stable. Further contrasts were completed to examine the slope differences between Aß+ Progressor and Aß+ Stable, Aß+ Stable and Aß– Progressor, and between Aß+ Progressor and Aß– Progressor. Other variables of interest also included follow up time and group status. The model also included vascular risk factors, age at baseline, sex, education, and APOE4 status as covariates. The categorical variable APOE4 was used to contrast subjects with one or two APOE ɛ4 alleles against those with zero. Participant ID was included as a categorical random effect.

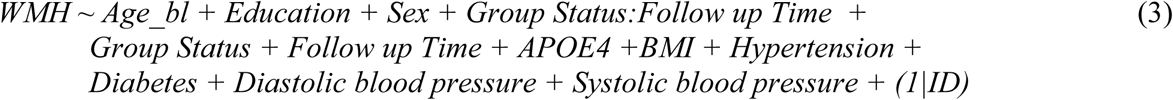

All continuous longitudinal values were z-scored within the population prior to the regression analyses. All statistical analyses were performed using MATLAB version 2021a. To complete the baseline analysis, we used function fitlm and for longitudinal assessments we used fitlme.

#### 2.6.4 ROC curve analysis

To further verify whether WMH progression impacts conversion to dementia in MCI individuals on the AD trajectory (i.e., Aß+ MCI populations), we calculated and used subject specific WMH progression slopes to distinguish between Aß+ Progressor and Aß+ Stable participants. WMH progression slopes were calculated by regressing the longitudinal WMH volumes (global and regional) against follow-up times as well as an intercept term (using regress function). We then applied receiver operating characteristic (ROC) curve analysis for total and regional slopes to differentiate Aß+ Progressor and Aß+ Stable participants. We performed the ROC analysis separately for groups with baseline CDR-SB values of 0.5, 1, 1.5, and 2, since individuals with different baseline CDR-SB values are at different stages of cognitive decline with different rates of conversion (i.e. MCI individuals with higher baseline CDR-SB values are closer to a threshold of dementia diagnosis), creating an unaddressed confound if assessed together We report area under the curve (AUC), a measure of separability between the two groups, for each region and total WMH.

## 3 Results

### 3.1 Baseline Demographics and Cognitive Scores

Table 1 presents demographic information and clinical characteristics of the participants included in the study. Aß– Stable was significantly younger than both Aß+ Progressor (x^2^= -3.48, *p<*.001) and Aß+ Stable (x^2^= -4.09, *p<*.001). The proportion of people with APOE ε4+ significantly differed between all groups except Aß– Stable and Aß– Progressor (x^2^ belongs to [12-122], *p*<.001), with Aß+ Progressor having the highest proportion of people with APOE ε4+, followed by Aß+ Stable, Aß– Stable, and lastly, Aß– Progressor. Years of education and proportion of male to females did not differ between any of the groups. All groups had significantly different Alzheimer’s Disease Assessment Scale-13 (ADAS-13) scores except for both Aß– and Aß+ Progressors, and Aß– Progressor and Aß+ Stable (*t* belongs to [4.43–21.12], *p*<.001). Aß+ Progressor exhibiting the lowest cognitive performance, followed by Aß– Progressor, then Aß+ Stable, and lastly, Aß– Stable with the best performance. For CDR-SB scores, Aß+ Progressor significantly differed from Aß+ and Aß-Stable (*t* belongs to [7.71-7.86], *p*<.001). Body mass index (BMI) was significantly lower in Aß+ progressors than Aß– Stable (*t*= -3.97, *p<*.001). As for vascular risk factors, hypertension, diabetes, and systolic and diastolic blood pressure were not significantly different between groups.

### 3.2 Baseline Assessments

Figure 1 shows boxplots of baseline total WMH load and separately for each lobe for all groups. Table 2 summarizes the linear regression model results for baseline WMHs for all groups across all regions. Both Aß+ groups exhibited greater WMH burden than the Aß– stable group (Aß+ progressor: *t* belongs to [2.97–3.73], *p*<.001, and Aß+ stable: *t* belongs to [2.73–3.94], *p*<.01) at all regions of interest.

**Figure 1:**
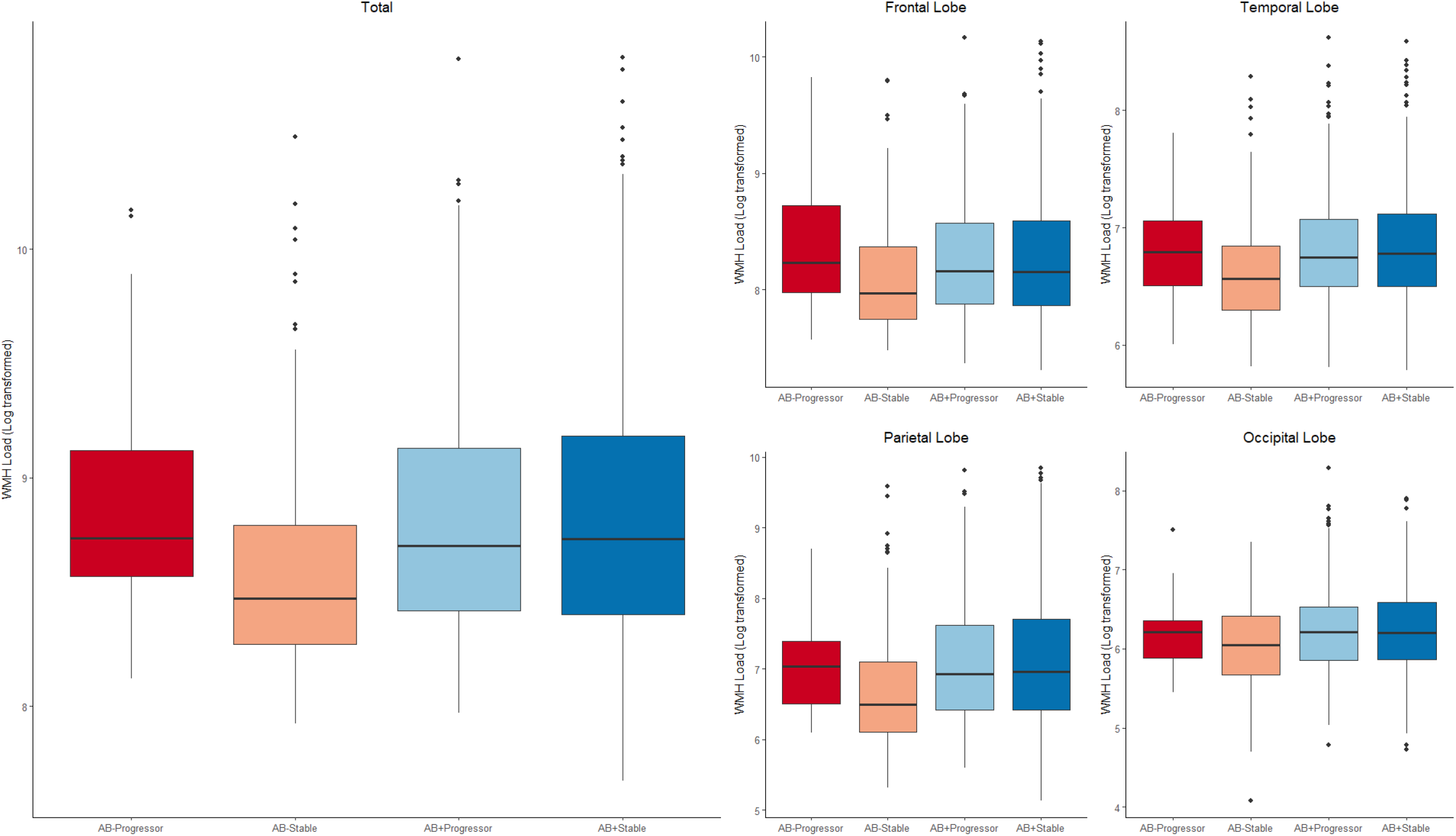
Boxplots showing baseline WMH distributions (log transformed) across diagnostic groups for each lobe. *Notes:* baseline WMH distributions (log transformed) across diagnostic groups for each lobe. The first and second rows show the log transformed WMH loads for each group by lobe. Amyloid positive (Aβ+) progressor, amyloid negative (Aβ–) progressor, amyloid positive (Aβ+) stable, and amyloid negative (Aβ–) stable. WMH = white matter hyperintensity

**Table 2:** Linear regression model results showing baseline differences between Aß+ Progressor, Aß– Progressor, Aß+ Stable, and Aß– Stable

|  | Total |  | Frontal |  | Temporal |  | Parietal |  | Occipital |  |
| --- | --- | --- | --- | --- | --- | --- | --- | --- | --- | --- |
|  | T stat | P | T stat | P | T stat | P | T stat | P | T stat | P |
| **WMH** |  |  |  |  |  |  |  |  |  |  |
| Aβ- Stable vs Aβ- Progressor | 1.39 | =.164 | 1.80 | =.072 | 1.04 | =.299 | 1.09 | =.276 | 0.75 | =.454 |
| Aβ- Stable vs Aβ+ Stable | 3.75 | <.001* | 3.07 | =.002* | 3.88 | <.001* | 3.94 | <.001* | 2.73 | <.001* |
| Aβ- Stable vs Aβ+ Progressor | 3.59 | <.001* | 2.97 | =.003* | 3.46 | <.001* | 3.73 | <.001* | 3.04 | <.001* |
| Aβ+ Stable vs Aβ+ Progressor | 0.06 | =.800 | 0.07 | =.795 | 0.008 | =.976 | 0.05 | =.814 | 0.49 | =.480 |
| Aβ+ Stable vs Aβ- Progressor | 0.03 | =.867 | 0.28 | =.595 | 0.33 | =.563 | 0.30 | =.581 | 0.15 | =.695 |
| Aβ- Progressor vs Aβ+ Progressor | 0.07 | =.793 | 0.18 | =.673 | 1.06 | =.303 | 0.39 | =.530 | 0.42 | =.517 |

Notes: \* Represents results that are significant when uncorrected and bolded values represent those that remained significant after correction for multiple comparisons.

### 3.3 Longitudinal Assessments

Figure 2 shows the longitudinal WMH distributions separately for each lobe for all groups. Table 3 summarizes the results of the longitudinal linear mixed effects models for all groups contrasted against Aß– stable across all WMH regions. Follow-up time was associated with a significant increase in WMH load in Aß– stable (*t* belongs to [7.15–21.33], *p*<.001). Aß– progressors (*t* belongs to [4.45–5.22], *p*<.001) had increased WMH load change over time compared to AB– stable at all regions except temporal. Aß+ stable (*t* belongs to [6.12–9.30], *p*<.001), and Aß+ progressors (*t* belongs to [10.77–19.17], *p*<.001) had significantly increased WMH load change over time in all regions compared to Aß– stable group. Aß+ progressors (*t* belongs to [3.87–11.01], *p*<.001) had significantly increased WMH load change over time in all regions compared to Aß+ stable group. Aß– progressors (*t*=2.89, *p*=.004) had increased WMH load change over time compared to Aß+ stable group at only the occipital region. Aß+ progressors (*t* belongs to [5.07– 6.45], *p*<.02) had significantly increased WMH load change over time in all regions compared to Aß-progressor group at total, frontal, and parietal regions only. That is, compared to Aß– stable, increasingly steeper change over time was observed in Aß– progressors, then Aß+ stable, then to Aß+ progressors. To ensure that the minimum duration of follow-up time does not impact these results, the models were repeated including only participants with a minimum of one, two, and three years of follow-up, yielding similar results in terms of effect size and significance.

**Figure 2:**
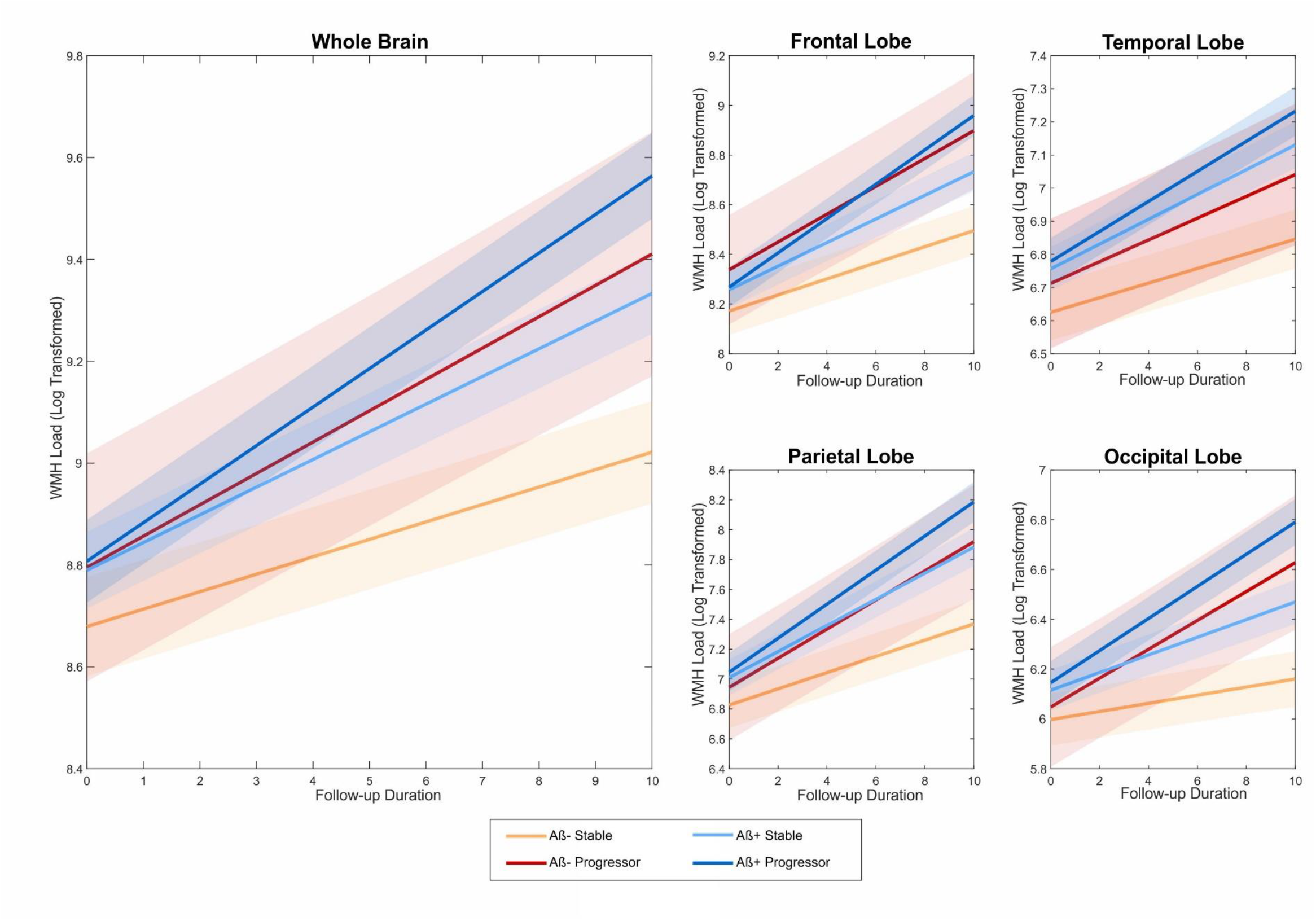
Longitudinal change in total and regional WMH volume by group *Notes:* Longitudinal WMH distributions (log transformed) across diagnostic groups for each lobe. The first and second rows show the log transformed WMH loads for each group by lobe. Amyloid positive (Aβ+) progressor, amyloid negative (Aβ–) progressor, amyloid positive (Aβ+) stable, and amyloid negative (Aβ–) stable. WMH = white matter hyperintensity

**Table 3:** Linear mixed model results showing longitudinal slope differences between Aß+ Progressor, Aß– Progressor, Aß+ Stable, and Aß– Stable

|  | Total |  | Frontal |  | Temporal |  | Parietal |  | Occipital |  |
| --- | --- | --- | --- | --- | --- | --- | --- | --- | --- | --- |
|  | *T* stat | *P* | *T* stat | *P* | *T* stat | *P* | *T* stat | *P* | *T* stat | *P* |
| **WMH** |  |  |  |  |  |  |  |  |  |  |
| FollowUpTime | 21.33 | <.001* | 21.27 | <.001* | 13.72 | <.001* | 20.88 | <.001* | 7.15 | <.001* |
| Aβ- Stable vs Aβ- Progressor : FollowUpTime | 4.87 | <.001* | 4.45 | <.001* | 1.93 | =.054 | 4.76 | <.001* | 5.22 | <.001* |
| Aβ- Stable vs Aβ+ Stable : FollowUpTime | 9.17 | <.001* | 7.27 | <.001* | 7.00 | <.001* | 9.30 | <.001* | 6.12 | <.001* |
| Aβ- Stable vs Aβ+ Progressor : FollowUpTime | 19.17 | <.001* | 17.95 | <.001* | 10.77 | <.001* | 17.07 | <.001* | 15.67 | <.001* |
| Aβ+ Stable vs Aβ+ Progressor : FollowUpTime | 10.30 | <.001* | 11.01 | <.001* | 3.87 | <.001* | 7.97 | <.001* | 9.89 | <.001* |
| Aβ+ Stable vs Aβ- Progressor : FollowUpTime | 1.30 | .193 | 1.62 | .105 | -.80 | .421 | 1.13 | .259 | 2.89 | .004* |
| Aβ- Progressor vs Aβ+ Progressor : FollowUpTime | 6.45 | .011* | 6.20 | .012* | 5.07 | .020* | 3.41 | .065 | 0.67 | .412 |

Notes: \* Represents results that are significant when uncorrected and bolded values represent those that remained significant after correction for multiple comparisons.

### 3.4 Vascular Risk factors

At baseline assessment, BMI and systolic and diastolic blood pressure were not significantly associated with WMHs at any region. Diabetes was observed to be a significantly associated with WMH only in the occipital region (*t*=-2.12, *p*=.03) showing no relationship in the other regions. Hypertension was found to be significantly associated with WMH in total volume (*t*=3.38, *p*<.001) and all regions (*t* belongs to [2.14–3.70], *p*<.035) except occipital.

At longitudinal assessment, BMI, systolic and diastolic blood pressure, and diabetes were not significantly associated with WMHs at any region (*p* > 0.05). Hypertension was found to be significantly associated with WMH in total volume (*t*=4.00, *p*<.001) and all regions (*t* belongs to [2.50–4.33], *p*<.015) except occipital. Although not significant, the strongest association between WMHs and diabetes was observed in the occipital region (*t*=-1.90, *p*=.056).

### 3.4 ROC for Classification: Amyloid Positive Converters vs Non-Converters

To distinguish between Aß+ progressors and Aß+ stable based on the longitudinal slope of WMH, we compared the predictive value of total and regional WMHs for each baseline CDR-SB value (0.5, 1, 1.5, and 2). As expected, conversion ratios were very different across groups with different baseline CDR-SB values. For baseline CDR-SB value of 0.5 (*N* = 111), conversion ratio was observed at 0.1875. For CDR-SB value of 1 (*N* = 118), 1.5 (*N* = 109), and 2 (*N* = 85), conversion ratios were observed at 0.3364, 0.4455, and 0.5467, respectively. ROC curve analyses showed that WMH slopes were able to differentiate between stable and progressor individuals. Classification performance was highest for the group with baseline CDR-SB value of 1, with best differentiation seen for the frontal (AUC = 0.71, Fig. 3) and total WMHs (AUC = 0.72, Fig. 3). Overall, total and frontal WMH slopes were very good differentiators of conversion, suggesting that WMH progression is a contributing factor to AD conversion, particularly in individuals in the initial stages of cognitive impairment (i.e., CDR-SB<2).

**Figure 3:**
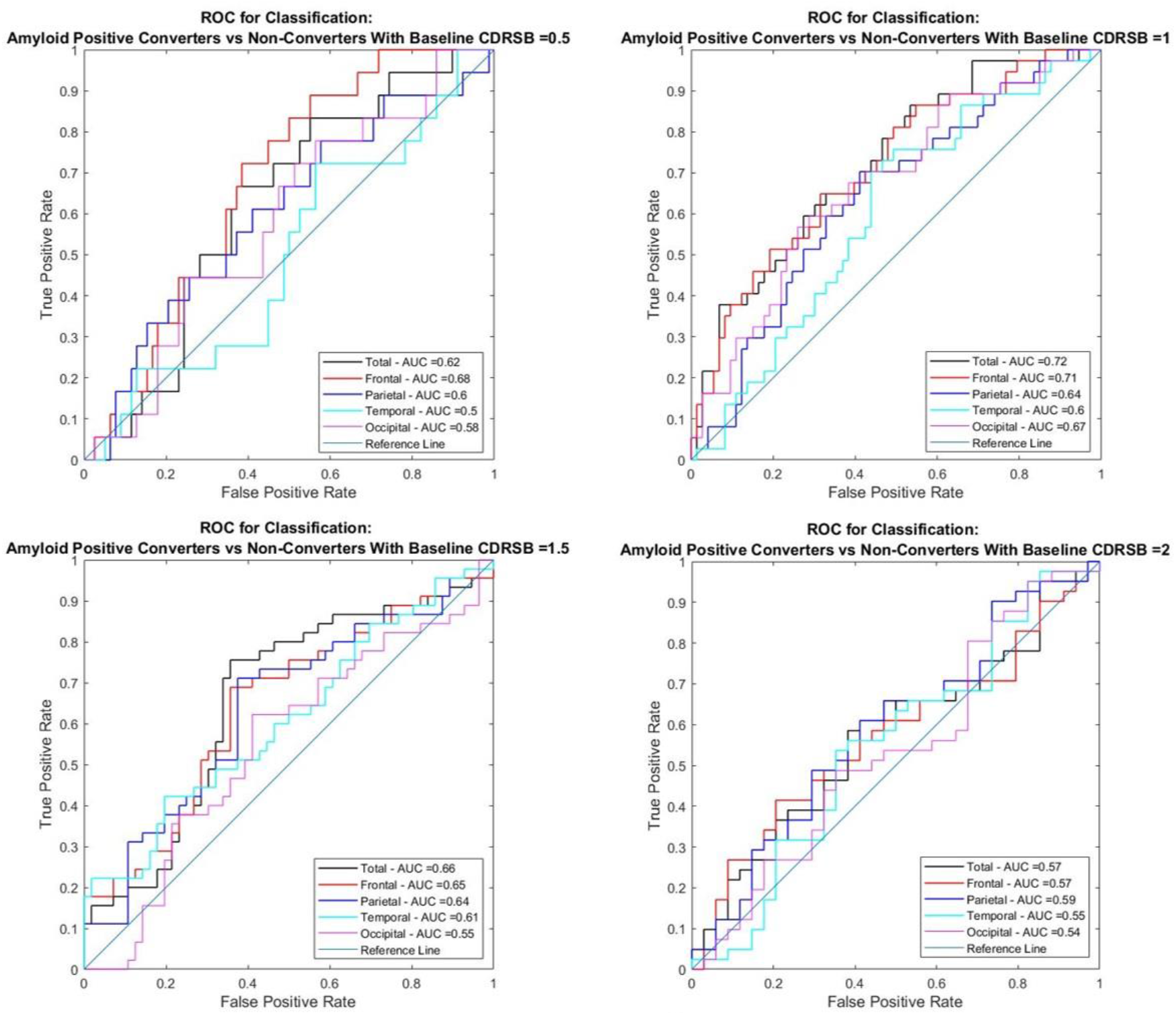
Receiver operating characteristics (ROC) analysis to compare the area under the curve (AUC) of Amyloid Positive Converters vs Non-Converters *Notes:* Receiver operating characteristics (ROC) analysis to compare the area under the curve (AUC) of Amyloid Positive Converters vs Non-Converters at 0.5,1,1.5 and 2 CDR-SB scores for regional and total WMH. CDR-SB = Clinical Dementia Rating – Sum of Boxes. WMH = white matter hyperintensity

## 4 Discussion

Previous research has identified an association between MCI and WMH load, with pMCI showing increased WMH loads compared to sMCI. This association, however, has not been examined in studies with long follow-ups to determine whether rate of WMH load change over time differs between the groups. Findings have also yet to explore whether amyloid positivity or negativity influences rate of change in WMH burden in progressive and stable MCI. The present study investigated regional WMH accumulation at baseline and longitudinally in four subtypes of people with MCI: Aß+ progressors, Aß+ stable, Aß– progressors, and Aß– stable. We have 4 main findings: 1) WMH burden was related to progression to both AD and dementia (i.e., associated with increased rate of change in both Aß+ progressors and Aß– progressors, 2) Aß+ positive individuals have increased WMH progression compared to Aß– individuals, 3) hypertension was significantly associated with baseline and longitudinal progression of WMH burden in total volume and all regions except occipital, and 4) slope of change in frontal and total WMHs can differentiate between Aß+ stable and progressor individuals with similar baseline cognitive status (CDR-SB), particularly in earlier stages of cognitive decline

We observed that Aß– stable had lower baseline WMHs compared to both Aß+ progressors and Aß+ stable across all regions. In line with previous reports, which had a shorter follow-up period, we found a disproportionately greater prevalence of WMHs in Aß+ progressors with increased rate of progression from MCI to dementia (Dadar et al., 2019). When examining the longitudinal data, we found that increased follow-up time was associated with increased WMH in all regions. In the present study, when examining change over time, compared to Aß– stable, all groups had significantly steeper change in WMH burden. The largest change (in slope) was observed for Aβ+ progressors compared to Aß– stable. Compared to positive stable, negative progressors only had steeper slopes (i.e., increased change in WMHs over time) in the occipital region. Posterior WMH accumulation has previously been associated with AD-associated degenerative mechanisms (Brickman, 2013; Habes et al., 2018; McAleese et al., 2021). Thus, although these amyloid negative individuals are progressing to non-AD dementia, they are exhibiting some WMH burden that resembles the pattern seen in individuals with AD.

In such cases, accumulation of WMH in the occipital region is an indicator of dementia conversion for amyloid negative individuals. Positive progressors exhibited a steeper slope over time than negative progressors (only in total volume, frontal, and temporal regions). This difference between negative and positive progressors suggests that WMH rate of change in these areas in people who develop dementia may be partly accounted for by amyloid positivity. This interpretation is further supported by the positive progressors exhibiting increased rate of WMH change compared to positive stable in the same regions as negative progressors (total, frontal, and temporal) but also in the parietal and occipital; indicating that the parietal and occipital changes might be associated with progression to dementia, whereas the total, frontal, and temporal WMHs may be associated with other factors (e.g., amyloid or vascular risk factors). This interpretation coincides with previous research suggesting that frontal white matter lesions are partly associated with small vessel disease (McAleese et al., 2021). These findings suggest that an increase in widespread WMH pathology occurs in MCI who progress to dementia. More specifically, regardless of amyloid status, older adults that convert to dementia have more WMH progression than those who remain stable.

Overall, our results highlight that regardless of amyloid status, older adults that convert to dementia have more WMH progression than those who remain stable. Thus, WMH progression is a contributing factor to the conversion to dementia both in Aß– and Aß+ older adults with MCI. These findings indicate that WMHs are important components for progression to dementia/AD. Additionally, Aß+ MCIs exhibited more WMH progression than Aß– MCIs. Therefore, our finding suggests that amyloid positivity is related to rate of progression of WMH across brain regions and at least a portion of WMH progression can be explained by amyloid positivity.

In addition to examining the relationship between WMH progression and amyloid status, we also examined the relationship between WMH burden and vascular risk factors as the direct underlying mechanism involved in WMH accumulation. The importance of examining these factors is that vascular risk factors are potentially preventable/treatable and with appropriate intervention, WMH progression might be controlled, which in turn can slow down the progression to dementia. To examine the association with vascular factors, diabetes, systolic and diastolic blood pressure, BMI, and hypertension were added to the baseline and longitudinal models. Our findings suggest that hypertension is strongly associated with both baseline and rate of accumulation of total and frontal WMHs. In this sample, approximately 48% of participants had hypertension, which allows for stronger conclusions regarding the influence of hypertension on WMH burden. This finding follows past research indicating the association between small vessel disease and anterior WMH burden (McAleese et al., 2021). Given that hypertension is a modifiable condition, it may be possible to prevent some of the WMH accumulation through blood pressure management. Prevention of WMH burden may in turn mitigate or slow progression to dementia (Kjeldsen et al., 2018; Williamson et al., 2019). Diabetes showed to be marginally associated with baseline occipital WMH burden which was no longer significant in the longitudinal findings when including other risk factors as covariates in the models. These limited findings are likely because the few participants identified as having diabetes (only 69 out of the entire 820 participants included). Therefore, these results need to be replicated in a larger sample of diabetic patients in order to make generalizable conclusions on the influence of diabetes on WMH burden in people with MCI. BMI and systolic and diastolic blood pressure were not significantly associated with baseline or longitudinal WMH accumulation. Other factors such as waist-to-hip ratio, which has previously shown to be associated with WMH burden (Veldsman et al., 2020), may be a more sensitive risk factor that should be examined in future MCI studies.

The ROC analysis (Figure 3) showed that total and frontal longitudinal WMH slopes at baseline CDR-SB score of 1 outperformed other regions and baseline CDR-SB scores at accurately classifying Aβ+ pMCI from Aβ+ sMCI (Total AUC = 0.72; Frontal AUC = 0.71). While previous research has shown that most individuals with Aβ positivity will eventually develop AD (Sperling et al., 2014), screening based on Aβ positivity generally leads to the inclusion of a considerable proportion of stable participants (Ossenkoppele et al., 2015). Our findings suggest that WMHs can improve classification of Aβ+ pMCI vs Aβ+ sMCI with relatively high accuracy (AUC = 0.72, Fig 3). These findings suggest that total and regional WMHs can aid in future research and clinical settings to assess trajectories of Aβ+ MCI patients on the AD continuum.

There are limitations to the present study that future research should explore. First, participants in the ADNI dataset have relatively high education levels and socioeconomic status. While our models included years of education as a covariate, future research should aim to examine the influence of education levels on WMHs in more representative Aβ negative and positive pMCI and sMCI populations, to determine whether high education levels are protective against WMH progression and the related conversion to dementia. Furthermore, the Aß– Progressor group had a smaller sample size compared to the other groups, limiting the statistical power and generalizability of its results. Although linear mixed models are robust with regards to unbalanced data and optimally use all information together without reducing power, a larger sample is warranted and would make the results comparing the baseline and longitudinal data involving Aß– Progressor individuals more conclusive.

The current study observed that WMH accumulation is an important factor in MCI progression to dementia for both Aß+ and Aß– individuals. This association was strongest in those who were Aß+, indicating that WMH accumulation is, in part, related to amyloid deposition. Hypertension was also found to be associated with increased WMH burden and rate of change in total volume and all regions except occipital, suggesting that progression from MCI to dementia may be mitigated through hypertension prevention and treatment.

## Data Availability

All data produced are available online at Alzheimers Disease
Neuroimaging Initiative (ADNI) database (adni.loni.usc.edu)

## Acknowledgments

Data collection and sharing for this project was funded by the Alzheimer’s Disease Neuroimaging Initiative (ADNI) (National Institutes of Health Grant U01 AG024904) and DOD ADNI (Department of Defense award number W81XWH-12-2-0012). ADNI is funded by the National Institute on Aging, the National Institute of Biomedical Imaging and Bioengineering, and through generous contributions from the following: AbbVie, Alzheimer’s Association; Alzheimer’s Drug Discovery Foundation; Araclon Biotech; BioClinica, Inc.; Biogen; Bristol-Myers Squibb Company; CereSpir, Inc.; Cogstate; Eisai Inc.; Elan Pharmaceuticals, Inc.; Eli Lilly and Company; EuroImmun; F. Hoffmann-La Roche Ltd and its affiliated company Genentech, Inc.; Fujirebio; GE Healthcare; IXICO Ltd.; Janssen Alzheimer Immunotherapy Research & Development, LLC.; Johnson & Johnson Pharmaceutical Research & Development LLC.; Lumosity; Lundbeck; Merck & Co., Inc.; Meso Scale Diagnostics, LLC.; NeuroRx Research; Neurotrack Technologies; Novartis Pharmaceuticals Corporation; Pfizer Inc.; Piramal Imaging; Servier; Takeda Pharmaceutical Company; and Transition Therapeutics. The Canadian Institutes of Health Research is providing funds to support ADNI clinical sites in Canada. Private sector contributions are facilitated by the Foundation for the National Institutes of Health (www.fnih.org). The grantee organization is the Northern California Institute for Research and Education, and the study is coordinated by the Alzheimer’s Therapeutic Research Institute at the University of Southern California. ADNI data are disseminated by the Laboratory for Neuro Imaging at the University of Southern California.

